# Assessment of Medication Adherence Among Heart Failure Patients in an Ambulatory Care Setting: A Prospective Observational Study

**DOI:** 10.64898/2026.01.23.26344702

**Authors:** Morouj Bakr, Loay Milibari, Sara Al Khansa, Ebtisam Alkhattaby, Sumaia Jambi, Ibrahim Jlaidan, Rana Almuwallad, Amjad Madkhali, Ashjan Altuwrqi, Mawaddah Alrhily, Sarah Alharbi, Sanaa AlSulami

## Abstract

**Background:** Medication non-adherence is a critical problem among patients with heart failure (HF). Current evidence has shown its association with increased morbidity, mortality and healthcare costs. Prescription discrepancy is a significant risk factor that can increase non-adherence and subsequently increases the risk of HF-related hospitalization and mortality. Current literature has not provided a clear understanding of the non-adherence problem or contributing factors among Saudi HF patients. Measuring the prevalence of non-adherence and its associated factors can direct clinicians to implement effective interventions to optimize pharmacotherapy benefits, and thus, improving outcomes.

**Aim:** To assess the Saudi Arabian population of HF patients for degree of adherence to their medications, the amount of medication discrepancies, and its association with re-hospitalization rate.

**Method:** A prospective observational study conducted at a tertiary care hospital on eligible HF patients attending the ambulatory clinic from July 2023 through April 2024. All patients were followed for six months. Primary outcomes were percentage of patients’ adherent to their medications and number and type of prescription discrepancies. Secondary outcomes were degree of health literacy, prevalence of non-adherence risk factors, and HF-related re-hospitalization rate. Adherence was measured utilizing the Eight-item Morisky Medication Adherence Scale (MMAS-8). Three-item Brief Health Literacy Screen (BHLS) was used to measure health literacy.

**Result:** A total of 202 patients were included in the study. Average age was 60 years, and 69% were males. For the primary outcomes, 43.5% of patients demonstrated high adherence, while 39.6% and 16.8% fell into medium and low adherence categories, respectively. Prescription discrepancies were identified in 51.5% of the patients. Causes of discrepancies ranged from patient generated, healthcare system generated, or multifactorial, generated by both, the patient and the system. Degree of health literacy was adequate in 23.8% of the patients, marginal and inadequate in 51.5% and 24.8%, respectively. Of potential non-adherence risk factors, polypharmacy, age ≥65 years, and marginal and inadequate health literacy, were the most common. HF-related re-hospitalization occurred in 18 patients, all of which were either non-adherent or had prescription discrepancies.

**Conclusion:** Among HF patients, medication non-adherence is a significant problem that is associated with increased morbidity and mortality. In our study, around half of the patients either experienced difficulties with adherence, prescription discrepancies, or both. Measuring the local prevalence of factors affecting non-adherence can be of use to identify strategies that suit our population the best, in order to mitigate their negative effect.

## 1. Background

***H****eart failure (HF)* is a complex clinical syndrome characterized by inability of the heart to sufficiently pump blood to meet the metabolic needs of the body. It is a growing public health problem in Saudi Arabia, where the incidence of HF is increasing. This is attributed to various factors such as aging population, high prevalence of risk factors like diabetes and hypertension, and lifestyle changes^1,2^.

*Medication non-adherence* is a critical problem among patients with HF and can be associated with increased morbidity, mortality, and healthcare costs^3^. Medication non-adherence is defined as the extent to which a patient’s medication-taking behavior differs from the agreed-upon plan of their healthcare provider^4,5^. Studies among HF patients in the U.S. showed that non-adherent patients have a mortality rate that is 1.5 to 2 times higher than adherent patients, and a hospitalization rate that is up to 4 times higher ^6,7^. Additionally, existing studies have shown that medication non-adherence is prevalent among cardiac patients in Saudi Arabia and has adverse impacts on patients’ health outcomes and healthcare resources ^8,9^. A prospective study was conducted to measure medication adherence by using Medication Event Monitoring System (MEMS) to predict rehospitalization and mortality. The results suggest that non-adherence contributes to higher rates of hospitalization^10^. Another cross-sectional study was conducted in tertiary care centers in Riyadh to define the prevalence of different levels of medication adherence among cardiac patients, and if there is any correlation between age and the level of medication adherence. It was found that 33.7% of participants had low adherence levels. Moreover, there was a positive correlation between age and the level of adherence. The authors of this study emphasized the need for further research to identify the causes and potential solutions for this issue among cardiac patients ^8^.

Measuring medication adherence can be challenging, however, there are several validated tools and scores that have been developed to assess medication adherence, including the Medication Event Monitoring System (MEMS), pill counts, pharmacy refill records, and self-reported measures such as the Morisky Medication Adherence Scale ^11,12,13^.

*Prescription discrepancy* is a significant factor that can affect medication non-adherence among HF patients ^5^. It is defined as any difference in medications, doses, or frequencies between the patient’s medication list and their electronic medical records leading to confusion for the patient ^14,15^. Examples of discrepancies include patient-generated, healthcare system-generated or both^15,16^. Such discrepancies can lead to non-adherence, which subsequently can increase the risk of HF-related hospitalization ^15.^

Despite the previous studies on this topic, there is still a gap in understanding the specific factors that contribute to nonadherence among HF patients in Saudi Arabia, as well as potential interventions to address this issue.

Therefore, it is crucial to identify the prevalence of the factors that contribute to medication non-adherence, such as, polypharmacy, advanced age, limited health literacy, side effects, or challenges with access to healthcare^17^.

This study aims to determine the rate of adherence at our HF ambulatory clinic, the prevalence of factors associated with medication non-adherence, the type and the number of medications discrepancies, if any, and their impact on the rate of hospitalization. The results of this study could provide valuable insights for healthcare providers and policymakers in Saudi Arabia to improve medication adherence rates among cardiac patients, reduce healthcare costs, and enhance patient outcomes.

## 2. Methods

### 2.1 Study design and participants

A prospective observational study conducted at King Abdulaziz Medical City, National Guard Health Affairs (KAMC-NGHA) in Jeddah, Saudi Arabia. The study aim is to assess medication adherence, medication discrepancies, and health literacy among HF patients in an ambulatory setting. Patients who are 18 years and old with confirmed diagnoses of HF and on guideline-directed medical therapy (GDMT) who are attending our HF ambulatory clinic between July 2023 and April 2024, were included. Patients following through HF telehealth clinic were excluded.

### 2.2 Data collection and follow up

Following obtaining Institutional Review Board (IRB) approval from the King Abdullah International Medical Research Center (KAIMRC), the eligible HF patients were first assessed by a HF specialist. During this visit, the specialist evaluated each patient’s clinical status, reviewed their HF medications, and identified any medication discrepancies, non-adherence, and factors associated with non-adherence. This information was documented in the patients’ electronic medical records. Following the initial assessment, a pharmacy resident reviewed each patient’s notes to document specific data points, including patient demographics and prescription discrepancies. All data were securely recorded in a password-protected data collection sheet to ensure confidentiality.

Each patient was interviewed by a HF-nurse specialist utilizing two questionnaires. A validated Arabic version of **Morisky Medication Adherence Scale (MMAS-8)** was used to assess adherence. It is a widely used eight-item questionnaire that evaluates adherence behaviors across multiple dimensions, which provides valuable insights into patient behaviors and identifies those who may require additional support to improve their medication adherence.

The other questionnaire was to assess health literacy. A validated Arabic version of the **Three-item Brief Health Literacy Screen (BHLS)** was used. BHLS is a brief tool designed to evaluate patients’ health literacy, especially beneficial for chronic disease populations such as HF patients.

The BHLS includes three questions assessing the frequency of needing help to read hospital materials, difficulty in learning about medical conditions due to reading challenges, and confidence in filling out forms independently.

After the initial clinic visit, all patients were followed up for six months to monitor for any HF clinical exacerbations that led to emergency department visits or hospitalizations.

### 2.3 Endpoints

The primary endpoints were the percentage of medication adherence among HF patients in an ambulatory care setting and the number and type of medication discrepancies, if any. Secondary endpoints included the degree of health literacy and its association with medication non-adherence, the factors associated with medications non-adherence, and the rate of HF rehospitalization associated with medication non-adherence and/or medication discrepancy.

### 2.4 Statistical Analysis

Descriptive statistics were used for the baseline characteristics. Categorical variables are presented as percentages while Continuous variables are presented as means and standard deviations. All Data were analyzed using Microsoft Office Excel 2021.

## 3. Results

### 3.1 Demographics

A total of 202 heart failure patients were included in the study. Baseline characteristics were summarized for the overall cohort and stratified by medication adherence status (**Table 1**), which was assessed at the baseline visit as a primary study outcome. The mean age was 60.4 ± 14.1 years and the mean weight was 80.5 ± 18.8 kg. The majority were male (140, 69.3%). HFrEF was the most common HF type (163/202, 80.7%); mean LVEF was 32.5 ± 9.9%, and the mean number of HF hospitalizations in the past 12 months was 0.74 ± 1.39.

**Table 1:** Baseline Characteristics of the Study Population Overall and Stratified by Medication Adherence Status at Baseline.

| Parameter | Total (n =202) | Adherent (n=88) | Non-adherent (n=114) |
| --- | --- | --- | --- |
| **Age (years), mean ± SD** | 60.4 ± 14.1 | 63.5 ± 11.2 | 58 ± 15.6 |
| **Gender, n.(%)** |  |  |  |
| Male | 140 (69.3) | 59 (67) | 81 (71) |
| Female | 62 (30.7) | 29 (32.9) | 33 (28.9) |
| **Weight (Kg), mean ± SD** | 80.6 ± 18.8 | 78.8 ± 19.3 | 81.9 ± 18.4 |
| **Types of HF, n.(%)** |  |  |  |
| HFrEF | 163 (80.7) | 70 (79.5) | 93 (81.5) |
| HFmrEF | 28 (13.8) | 13 (14.7) | 15 (13.1) |
| HFpEF | 11 (5.4) | 5 (5.6) | 6 (5.2) |
| **LVEF %, mean ± SD** | 32.5 ± 9.9 | 32.8 ± 9.3 | 32.3 ± 10.3 |
| **Number of HF hospitalization in the past 12 months, mean ± SD** | 0.78 ± 1.39 | 0.8 ± 1.6 | 0.7 ± 1.2 |

Data are presented as numbers (%) and mean ± SD

**Table 2** presents the baseline medications used by heart failure patients. Beta-blockers were used in all patients (100%). The most common additional therapies were SGLT2 Inhibitor (93.1%), Loop Diuretics (74.3%), and Mineralocorticoid Receptor Antagonists (77.2%), while among RAAS inhibitors ARNI was the most common (63.4%). In contrast, digoxin (14.4%), Isosorbide Dinitrate (16.8%), ACE inhibitors (15.3%), ARB (7.9%) and Ivabradine (5.9%) were less frequently used.

**Table 2:** Baseline Guideline-Directed Medical Therapy (GDMT)

| **Baseline Medications** | **Total (n =202)** | **Adherent (n=88)** | **Non-adherent (n=114)** |
| --- | --- | --- | --- |
| **Beta Blockers** | 202 (100) | 88 (100) | 114 (100) |
| **Digoxin** | 29 (14.4) | 17 (19.3) | 12 (10.5) |
| **Ivabradine** | 12 (5.9) | 4 (4.5) | 8 (7) |
| **RAAS inhibitors:** |  |  |  |
| • Angiotensin Converting Enzyme Inhibitor (ACEi) | 31 (15.3) | 10 (11.3) | 21 (18.4) |
| • Angiotensin Receptor Blocker (ARB) | 16 (7.9) | 10 (11.3) | 6 (5.3) |
| • Angiotensin Receptor Neprilysin Inhibitor (ARNI) | 128 (63.4) | 57 (64.7) | 71 (62.2) |
| **Diuretics:** |  |  |  |
| • Loop Diuretics* | 150 (74.3) | 68 (77.3) | 82 (71.9) |
| • Metolazone | 3 (1.5) | 1 (1.1%) | 2 (1.7) |
| **Mineralocorticoid Receptor Antagonists** | 156 (77.2) | 70 (79.5) | 86 (75.4) |
| **Isosorbide Dinitrate** | 34 (16.8) | 12 (13.6) | 22 (19.3) |
| **Hydralazine** | 22 (10.9) | 10 (11.3) | 12 (10.5) |
| **SGLT2 Inhibitor** | 188 (93.1) | 83 (94.3) | 105 (92.1) |

Data are presented as numbers (%).
\* Loop Diuretics: frusemide, bumetanide
SGLT2i: Sodium-Glucose Cotransporter – 2 Inhibitors.

### 3.2 Primary Outcomes

#### Medication Adherence Levels Among Heart Failure Patients

The pie chart in **Figure 1** illustrates the percentage of medication adherence among HF patients in an ambulatory care setting. Of the patients analyzed, 43.6% showed high adherence, 39.6% exhibited medium adherence, and 16.8% had low adherence based on the MMAS-8. These findings underscore the variability in adherence levels within this population.

**Figure 1.**
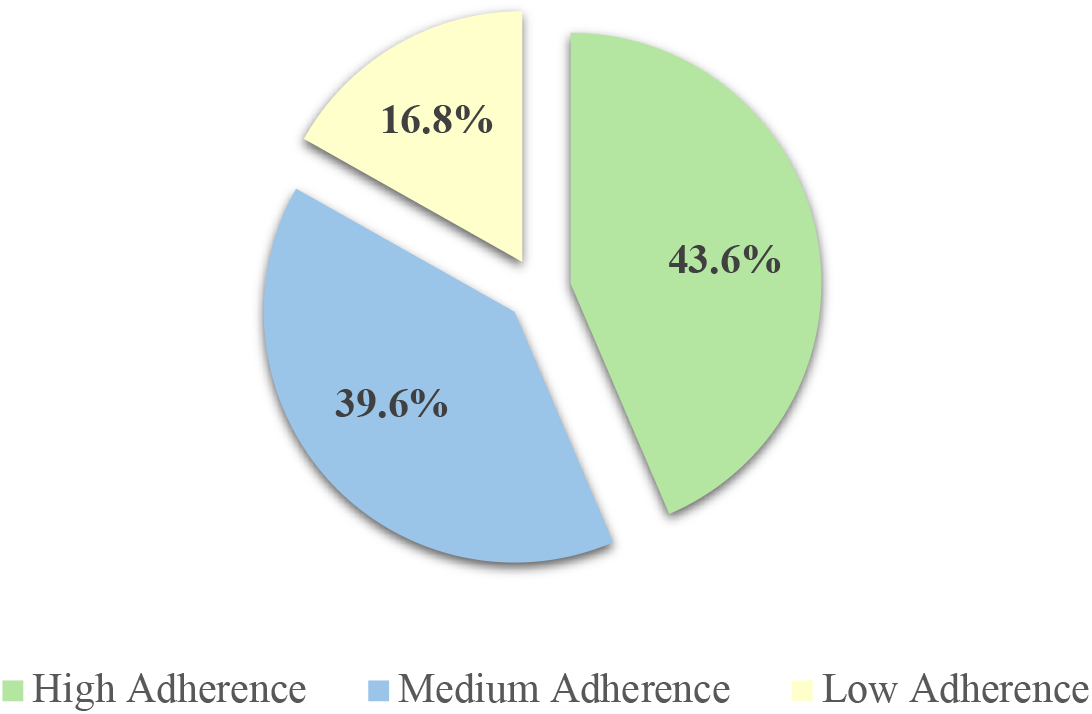
The Percentage of Medication Adherence Among HF Patients in an Ambulatory Care Setting

#### Types and Sources of Medication Discrepancies

A total of 178 medication discrepancies were identified in the study. These discrepancies were categorized based on their primary source into patient-generated, healthcare system-generated, or mixed (both patient and healthcare system-generated) discrepancies. Patient-generated discrepancies (PGD) were those arising from patient behavior or reporting and included medications prescribed but not taken by the patient, incorrect dosing, and deviations from the prescribed frequency of administration. Healthcare system-generated discrepancies (HSGD) originated from failures within healthcare processes or documentation, most notably the omission of active medications from the electronic medical record (EMR). Mixed discrepancies reflected contributions from both patient- and system-related factors and included medications prescribed by different hospitals or multiple prescribers, refill-related errors, and discrepancies related to different brand names. These mixed discrepancies commonly occurred during transitions of care and underscored the shared responsibility between patients and healthcare systems in maintaining accurate medication records.

As shown in **Figure 2**, PGD were the most prevalent, accounting for 70.2% (125 cases) of the total discrepancies. This was mainly due to patients taking their medications different than prescribed (different dosing, frequency or both) which was observed in 76 cases of loop diuretics, or not taking their prescribed medications at all as seen in 49 cases of SGLT2 inhibitors.

**Figure 2.**
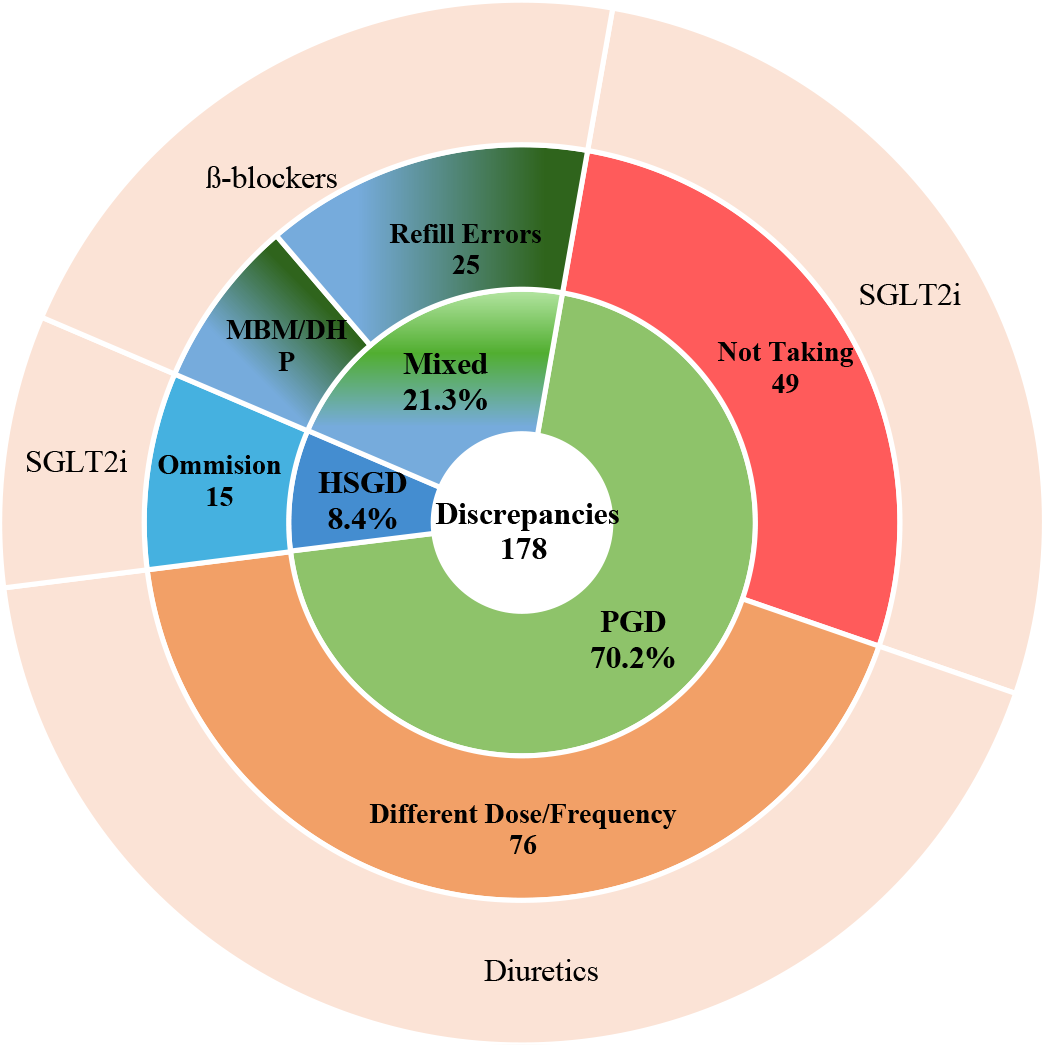
This chart provides a visual breakdown of the types of discrepancies, highlighting patient-related issues as the most significant source, followed by system-related and combined factors. Data are presented as numbers (%). Abbreviations: PGD: Patient-Generated Discrepancy, HSGD: HealthCare System –Generated Discrepancy, MBM: Multiple Brand name Medications, DHP: Different Hospital Prescribers, SGLT2i: Sodium-Glucose Cotransporter – 2 Inhibitors.

HSGD accounted for 8.4% of the total discrepancies, with the most notable issue being omissions from the electronic medical record (EMR). SGLT2 inhibitors were particularly impacted by these omissions (15 cases).

The remaining 21.3% of discrepancies were attributed to both patient and system generated factors. The most prominent issue in this group was refill errors, identified in 25 cases, predominantly involving beta-blockers. Discrepancies related to patients having medications from multiple prescribers and/or different brand names were also noted in 13 cases.

### 3.2. Secondary Outcomes Levels of Health Literacy

As part of the secondary outcomes, health literacy level was assessed using the BHLS. The results, as shown in **Figure 3**, revealed that nearly half of the study population (51.5%) had marginal health literacy. An additional 23.8% of patients were found to have adequate health literacy, while 24.8% had inadequate health literacy.

**Figure 3.**
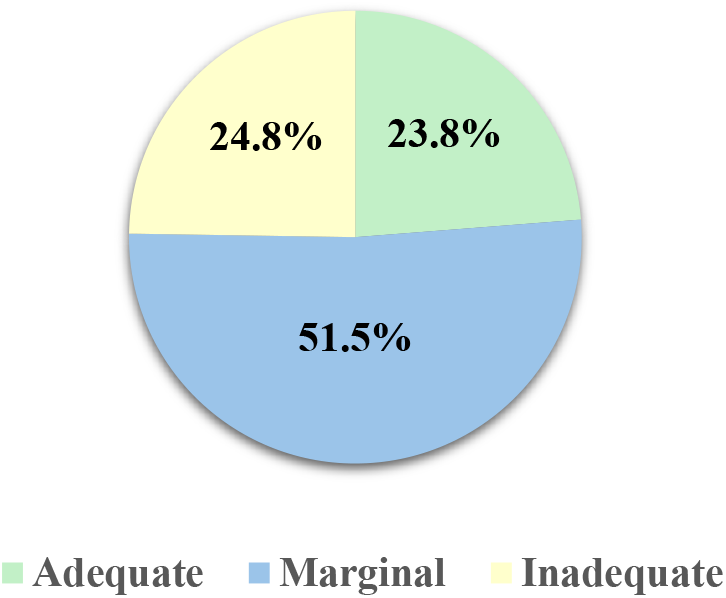
This pie chart represents the proportions of patients with varying levels of health literacy, highlighting marginal literacy as the most prevalent.

#### Factors Associated with Medication Non-Adherence

Multiple factors contributed to medication non-adherence among the study population (**Figure 4**). Polypharmacy was the most significant factor, affecting 97.37% of non-adherent patients. Marginal or inadequate health illiteracy was also prevalent, with 78.95% of non-adherent patients falling into this category. Older age (age ≥ 65 years) and adverse effects from medications were noted in 40.35% and 20.18% of non-adherent patients, respectively. Access to healthcare was the least common factor, affecting only 3.51% of the non-adherent group.

**Figure 4.**
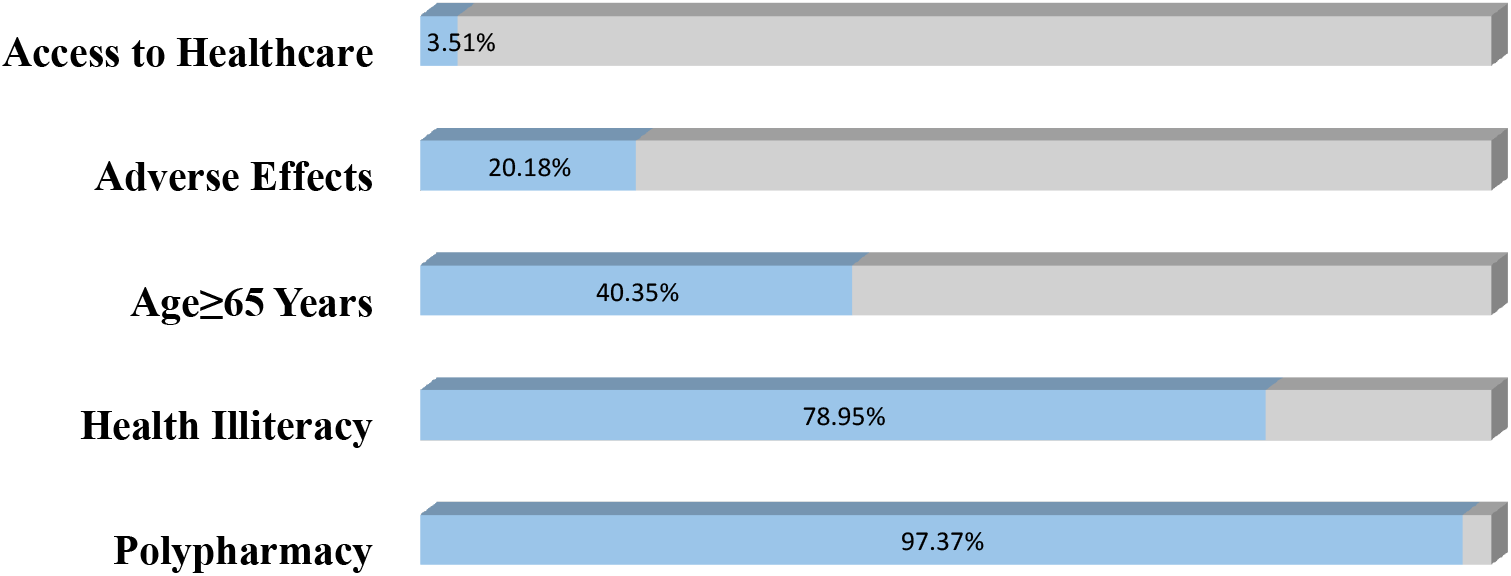
Prevalent factors associated with medication non-adherence: highlighting polypharmacy as the most common

#### Rate of Heart Failure Rehospitalization

At the six-month follow-up, 18 patients (8.9% of the study population) were rehospitalized. Of which, 13 patients (72.2%) were rehospitalized due to medication non-adherence, and 5 patients (27.7%) were rehospitalized due to medication discrepancies, as depicted in **Figure 5**. This breakdown highlights the significant role both medication discrepancies and non-adherence can play in heart failure rehospitalization rate.

**Figure 5.**
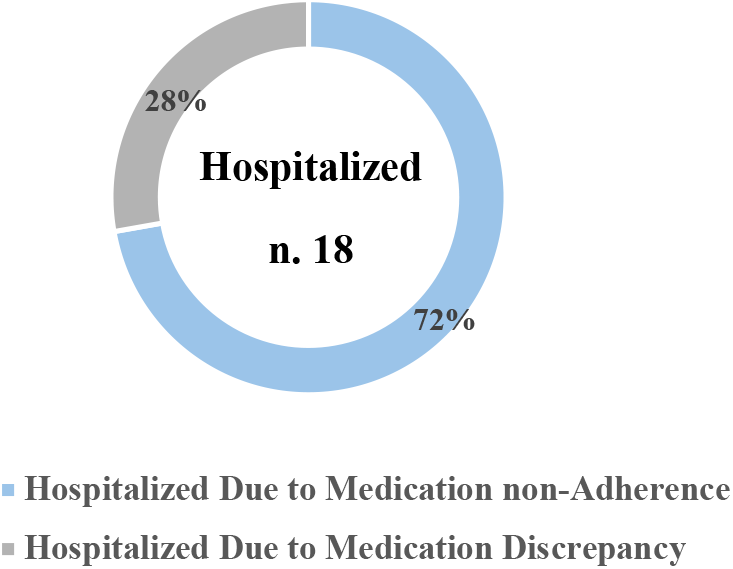
Rate of HF Rehospitalization and its association with Medications Non-Adherence or Discrepancy

## 4. Discussion

Our findings reveal significant challenges in medication adherence and highlight important considerations for improving heart failure management. The study found that only 43.5% of patients (n = 88) were adherent to their medications, while 39.6% (n=80) and 16.8% (n=34) fell into medium and low adherence categories, respectively. Our results are in line with the previous studies that were conducted in the Aseer region; they found 53.6% of the patients had poor medication adherence. However, the adherence rate for the patients’ medication was poor due to forgetfulness and was more common among single, unemployed patients with low income ^9,22^. This high rate of non-adherence is concerning, as poor medication adherence in heart failure patients has been associated with increased morbidity, mortality, and healthcare costs^9^.

The high prevalence of medication discrepancies, identified in 51.5% of patients, is alarming and suggests significant gaps in medication management. This rate is higher than previously reported in a study at King Fahad Medical City in Riyadh, which found discrepancies in 23.2% of cardiac patients^23^. The multifactorial nature of these discrepancies, involving both patient and healthcare system factors, highlights the complexity of the issue and the need for comprehensive strategies to address it.

Interestingly, our results showed that the mean age was similar between adherent (63.5 years) and non-adherent (58 years) patients. This suggests that age alone may not be a reliable predictor of adherence in this population, highlighting the need for a more nuanced approach to identifying patients at risk of non-adherence. This finding contrasts with some previous studies that have reported age as a significant factor in medication adherence^24^. However, the overall predominance of males in the study (about two-thirds of participants) raises questions about potential gender disparities in HF diagnosis or treatment in this setting, which may warrant further investigation. Moreover, medication nonadherence was numerically more prevalent among male patients in our cohort, similar to patterns described in a study conducted in the UAE ^25^.

The distribution of heart failure types was similar between adherent and non-adherent groups, with HFrEF being the most common (79.5% and 81.5%, respectively). This suggests that the type of heart failure may not significantly influence adherence patterns.

The health literacy assessment revealed that nearly half of the study population (51.5%) had marginal health literacy, with an additional 24.8% having inadequate health literacy. This finding is crucial, as low health literacy has been associated with poor health outcomes and increased healthcare utilization ^26^. The high prevalence of health illiteracy among non-adherent patients (78.95%) further emphasizes the importance of addressing this issue in interventions aimed at improving adherence. This is higher than rates reported in systematic review, which found inadequate health literacy in 39% of heart failure patients ^27^. Addressing health literacy should be a priority in efforts to improve medication management and overall patient care.

The identification of polypharmacy, older age, and health illiteracy as common risk factors for non-adherence provides potential valuable targets for intervention. These factors are consistent with existing literature on medication adherence in chronic diseases ^28,29^. While the relationship between age and adherence is complex, several studies have found that older age, particularly over 65, can be associated with lower medication adherence ^28,30^ and suggest the need for tailored approaches to support vulnerable patient groups.

The high prevalence of medication discrepancies in our study, particularly those originating from patient-related factors (70.2%), underscores the challenges in maintaining accurate medication lists and ensuring proper adherence. This finding aligns with previous research that has identified patient-related factors as significant contributors to medication discrepancies ^31^. The prominence of discrepancies related to SGLT2 inhibitors and diuretics is noteworthy, given their importance in heart failure management. The involvement of both patient and system factors in 21.3% of discrepancies, especially in refill errors/miscommunication, highlights the need for improved patient education and streamlined communication processes between healthcare providers and patients. The relatively lower proportion of purely system-generated discrepancies (8.4%) suggests that while electronic medical record systems have improved medication documentation, there is still room for enhancement, particularly in preventing omissions of critical medications like SGLT2 inhibitors. These findings emphasize the need for a multifaceted approach to reducing medication discrepancies, involving patient education, improved communication, and refined healthcare system processes ^23,32^.

The strong association between non-adherence or prescription discrepancies and rehospitalization is a critical finding. All 18 patients who experienced HF-related rehospitalization were either non-adherent or had prescription discrepancies. This underscores the potential impact of improved medication management on reducing healthcare utilization and improving patient outcomes.

Our study has several limitations. The single-center design and relatively small sample size of 202 patients may limit generalizability to the broader Saudi Arabian population with HF. Additionally, the use of self-reported health literacy measures (3-BHLS) may be subject to recall bias and social desirability bias. In addition, all patients were monitored for 6 months, which is relatively short period. However, we attempted to mitigate these limitations by using a validated health literacy scale, adherence scale and cross-referencing patient reports with prescription records and pill counts when possible.

Despite these limitations, our study has several strengths. The prospective design allowed us to capture real-time data on non-adherence and associated factors. We used a comprehensive approach to assess not only adherence but also health literacy, medication discrepancies, and rehospitalization rates, providing a holistic view of the challenges in heart failure management. Furthermore, our study is one of the few to examine these issues in a Saudi Arabian context, filling an important gap in the literature on heart failure management in this region. The six-month follow-up period provided valuable data on rehospitalization rates, allowing us to demonstrate the tangible clinical impact of non-adherence and medication discrepancies. This longitudinal aspect strengthens the clinical relevance of our findings. Lastly, our detailed analysis of medication types and discrepancies offers specific insights that can guide targeted interventions in clinical practice.

## 5. Conclusion

Among HF patients, medication non-adherence is a significant problem that is associated with increased morbidity and mortality. In our study, around half of the patients either experienced difficulties with adherence, prescription discrepancies, or both. Measuring the local prevalence of factors affecting non-adherence can be used to identify strategies that suit our population the best, in order to mitigate their negative effect.

## Data Availability

All data produced in the present work are contained in the manuscript

